# A Multimodal Foundation Model for Discovering Genetic Associations with Brain Imaging Phenotypes

**DOI:** 10.1101/2024.11.02.24316653

**Authors:** Diego Machado Reyes, Myson Burch, Laxmi Parida, Aritra Bose

**Affiliations:** Rensselaer Polytechnic Institute, Troy, NY; IBM Research, Yorktown Heights, NY

**Keywords:** GWAS, Foundation models, Constrative learning, Genomics, MRI, Neurological Disorders, AI

## Abstract

Due to the intricate etiology of neurological disorders, finding interpretable associations between multi-omics features can be challenging using standard approaches. We propose `COMICAL`, a contrastive learning approach leveraging multi-omics data to generate associations between genetic markers and brain imaging-derived phenotypes. `COMICAL` jointly learns omic representations utilizing transformer-based encoders with custom tokenizers. Our modality-agnostic approach uniquely identifies many-to-many associations via self-supervised learning schemes and cross-modal attention encoders. `COMICAL` discovered several significant associations between genetic markers and imaging-derived phenotypes for a variety of neurological disorders in the UK Biobank as well as predicting across diseases and unseen clinical outcomes from the learned representations. Source code of `COMICAL` along with pre-trained weights, enabling transfer learning is available at https://github.com/IBM/comical.

## 1 Introduction

Complex diseases such as neurological disorders are usually the result of the combined effect of multiple genes, commonly known as the polygenic effect, and multiple phenotypes or traits. These interactions are often induced by their symptoms or diagnosis. To obtain a holistic view of the disease, understand its origins, its mechanisms, causes, and consequences, it is necessary to study other modalities of data from omics studies such as genomics, proteomics, transcriptomics, radiomics, etc. An important challenge is to parse the multiomics data and find interpretable associations between them for accelerating discovery of therapeutics. Most studies that try to find associations between different omics with respect to a disease outcome tend to integrate data across multiple omics layers. One such approach involves co-mapping, where variables from two omic profiles are integrated and associated independently using regression-based methods [12]. Popular methods such as genome-wide association studies (GWAS), or quantitative expression trait loci (eQTL), are ways of associating genomics with other omics and understanding the impact of genetic variants on a target phenotype [4]. Some sophisticated machine learning methods have been proposed to predict and analyze neurological disorders [18, 20, 32], with limited methods in exploring multi-omics approaches and AI foundation models.

Recent advances in large language models or foundation models have shown that they can accurately predict the effects of missense variants [5, 8, 17], gene expression [2], and proteins [30]. These approaches performed representation learning on a specific omics data such as genomics, transcriptomics, proteomics, etc. and used these representations to understand genetic mechanisms impacting the phenotype. However, none of these approaches utilized recent advances in multimodal foundation models, such as vision language models, which propose a unified architecture of learning representations of image and text data [15] to integrate and associate multi-omics data. A previous study performed contrastive learning using self attention to classify multi-omics data using incomplete data but did not provide associations between multi-omics data and neither performed cross-disorder prediction [33]. Here, we propose `COMICAL` (contrastive multi-omics association learning), an adaptation of multimodal transformer CLIP (Contrastive Language-Image Pretraining) [21] to multi-omics analysis where it integrates two omics profiles to learn their representations and associate them. The training objective connects representations of one omics profile with the other and tries to discover “appropriate” vectors that capture one from the other using a contrastive loss function.

We evaluate `COMICAL` to find genotype-phenotype relationships in complex neurological diseases using genomic single nucleotide polymorphisms (SNPs) and radiomics or image-derived phenotypes (IDPs) from UK Biobank (UKB). `COMICAL` finds many-to-many associations between genetic variants associated with diseases such as Alzheimer’s Disease (AD), Attention-deficit/hyperactivity disorder (ADHD), Bipolar Disorder (BD), Cerebrovascular Disease (CBVD), Unipolar Depression (UD), Mood Disorder (MD), Multiple Sclerosis (MS), Parkinson’s Disease (PD), stroke, and Schizophrenia (SZ). `COMICAL` learns paired representations of IDPs and single nucleotide polymorphisms (SNPs) for these diseases using contrastive learning after tokenizing and encoding them using a scheme similar to CLIP [21]. `COMICAL` was able to accurately find the IDP and SNP pairs up to 100%, validated using an independent ENIGMA summary statistics. `COMICAL` also performs cross-disorder prediction and computes a novel multi-omics risk score computed from the joint learned representation of SNPs and IDPs. We show that `COMICAL`’s risk score is associated with polygenic risk scores from UKB.

Another key challenge in multi-omics analyses is the availability of annotated multi-omics data across modalities for complex diseases due to several privacy and technical reasons. Here, we make the pre-trained multimodal model weights available for SNP-IDP pairs, which can be used for fine-tuning tasks with independent data genomic and radiomics data. The many-to-many multi-omics associations produced as a result of this work along with the pre-trained models allow for accelerated discovery of multi-omics relationships through fine-tuning on genomic datasets and large biobanks.

## 2. Materials and Methods

### 2.1 Pretraining

CLIP [21] has shown to be an effective pre-training and learning strategy for image and text pairs, with efficient zero-shot transfer learning. `COMICAL` inherits the CLIP framework and pre-processes SNPs and IDPs, creating SNP-IDP pairs mediated by complex diseases listed above, and encodes them using transformer encoders as used in GPT-2 [22, 31]. From UKB [26], we obtain 40,426 samples with both SNPs and 154 IDPs of T1 structural brain MRI after quality control. We obtained 5,603 variants from the GWAS catalog [25] associated with the eight neurological diseases discussed above. After creating the pairs using top 1% SNPs in GWAS catalog (33 SNPs), we obtained 15,442,732 pairs as our pre-trained data. We split this into 70%, 20%, 10% scheme for training, validation, and test sets. For the IDP and SNP encoder, we use a 2-layer, 64-wide model with 4 attention heads and dimensions of 32 nodes in the feed-forward layers, with a learning rate of 1×10^−5^. The overview of `COMICAL` is shown in Figure 1 and the training and evaluation steps are outlined in Algorithm 1.

**Fig. 1:**
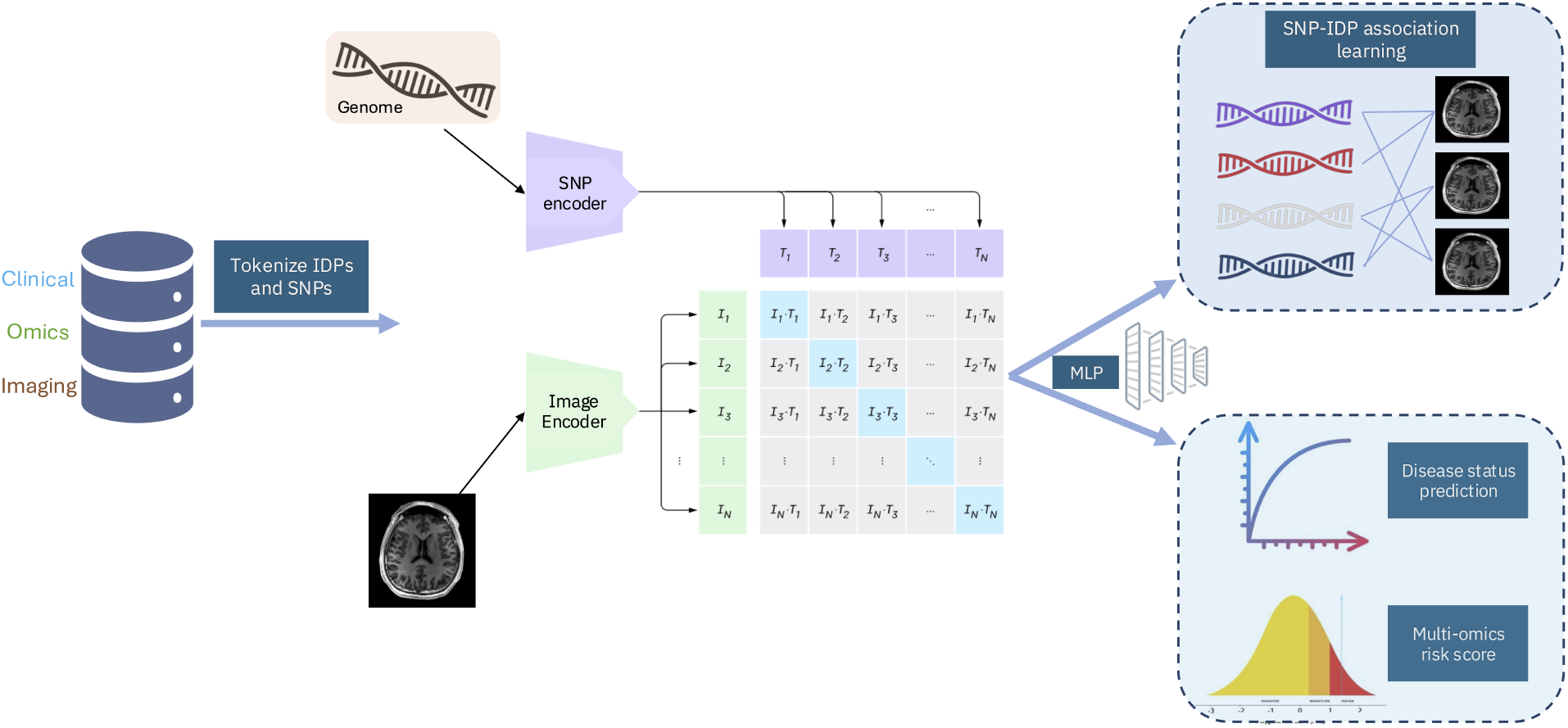
`COMICAL` inherits the framework of CLIP, tokenizes and encodes SNPs and IDPs using a transformer with self-attention masks and predicts the correct multi-omics pairings using a contrastive loss. In the evaluation phase, the learned encoder is used in a zero-shot linear classifier to predict the pairs.

#### Algorithm 1

`COMICAL`

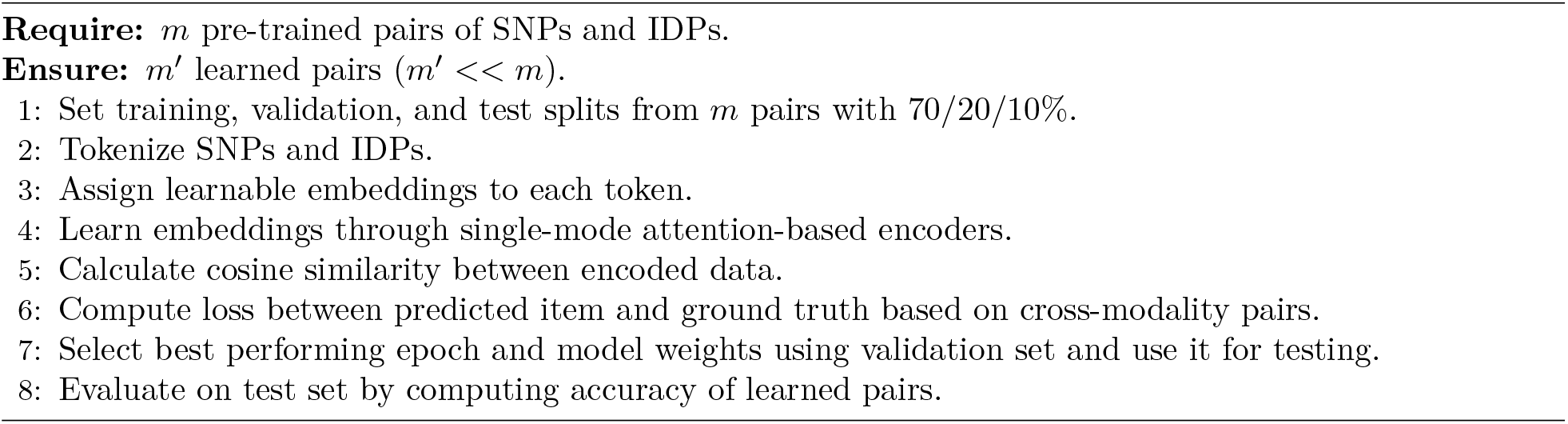

### 2.2 Multi-omics pairs

Pair-making is an essential step for `COMICAL` as this sets the ground truth for the model-predicted associations. Positive pairs between IDPs and SNPs were defined to perform contrastive training mediated by a set of eight diseases (AD, ADHD, BD, CBVD, MD, MS, PD, UD) as a proxy to establish the link between IDP and SNP. The key assumption behind pair making is that a relationship exists between an IDP and a SNP if both are related to the same disease. However, this is not always true, or it could be a weak relationship. Therefore, if the model consistently predicts two of them to be associated, then we can infer that there is, in fact, a strong relationship between them. This association learning is guided by the SNP and IDP values from all the samples.

Three main steps are taken to achieve this.

1. For each disease, create a set of SNPs which are associated with them from the GWAS catalog.
2. Perform a phenome-wide association study (PheWAS) using the `PheWAS` package in R [7], to map IDPs to diseases using covariates of age, sex, smoking status, height, BMI, and top ten genetic principal components (PCs). Create a set of one-to-one mapping between IDPs and diseases.
3. Expand the data so that each patient’s data becomes a record of SNPs and IDPs.
4. For each disease create data dictionaries relating for each IDP the SNPs that are associated with the same disease. Hence, making the pairs.

The pair-making process is outlined in Figure 2

**Fig. 2:**
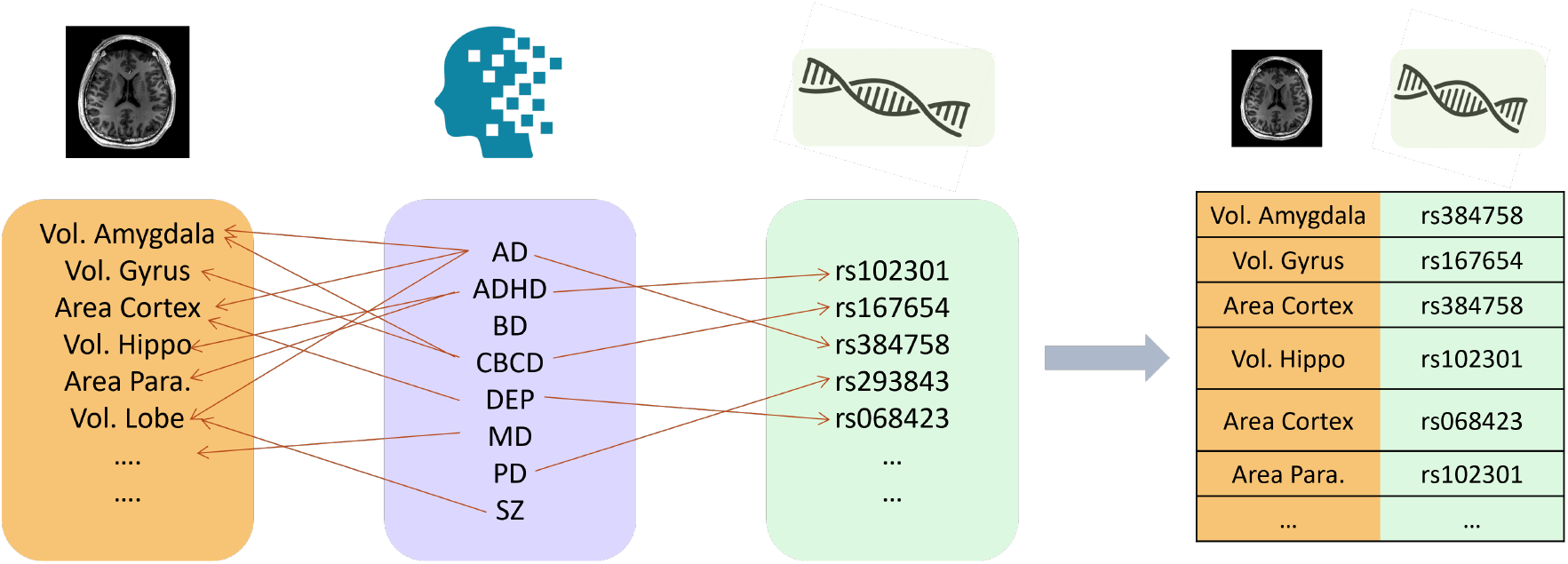
Pair-making process of `COMICAL` between SNPs and IDPs mediated by neurological diseases.

### 2.3 Tokenization

#### Imaging derived phenotypes

As IDPs are more closely related to tabular data than sequence data, we took an alternative approach to IDP tokenization using piecewise linear encodings, which are known to work well for numerical features in attention-based encoders [10]. Each IDP is first binarized and embeddings are computed from each sample based on their value relative to the rest. An *n*-dimensional (*n* samples) vector is built in which all the values are ones before the bin, where the value belongs, and all the following values are zeros (Figure 3a).

**Fig. 3:**
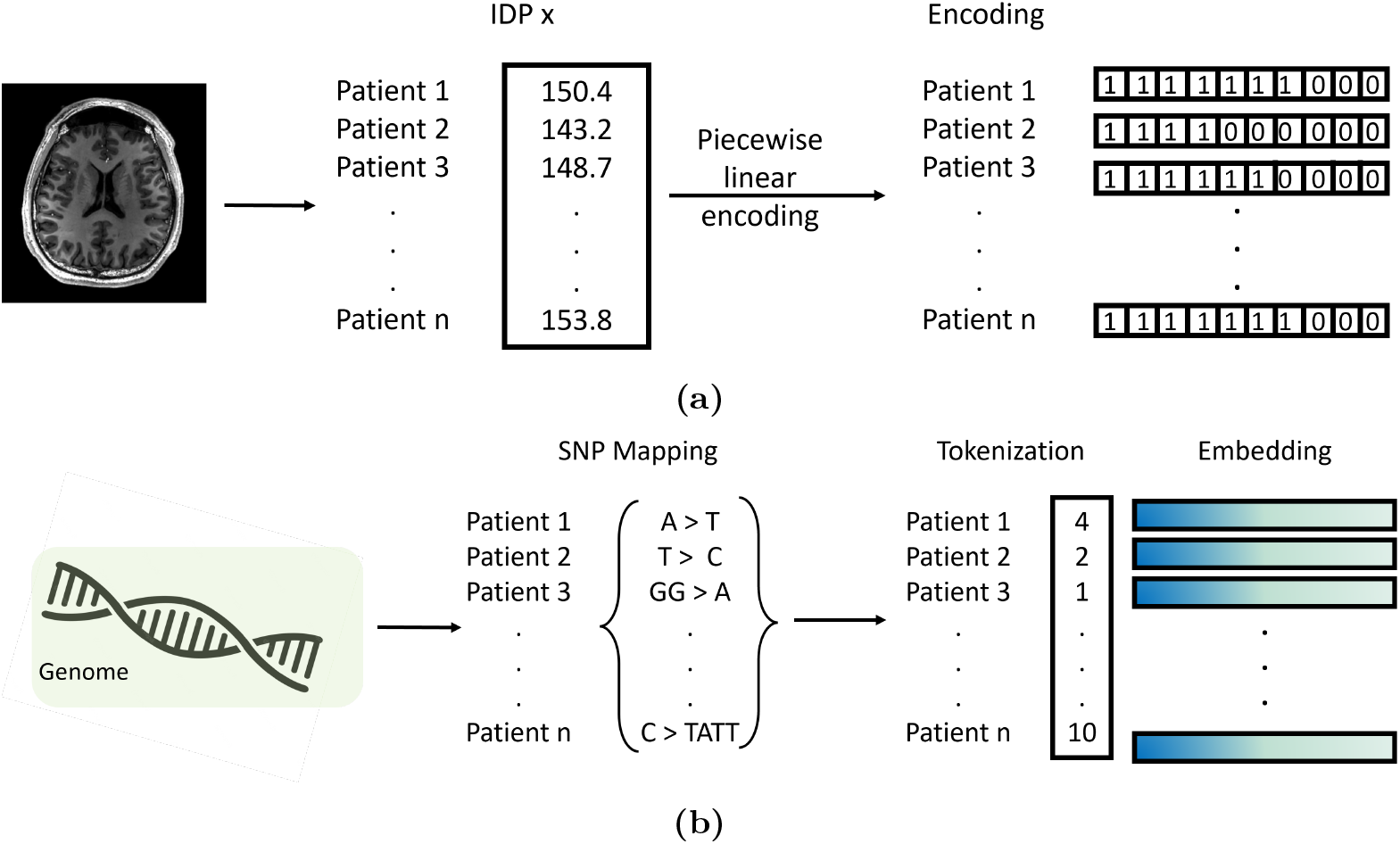
(a) Tokenization of IDPs using piecewise linear encoding (b) Tokenization of SNPs.

#### Genetic markers

The SNPs are tokenized for learning higher dimensional embeddings that can capture semantic relationship between them. We encoded the SNPs using the following process:

1. We created a dictionary (vocabulary of tokens) of all mutation types including heterozygous, homozygous, substitutions, insertions, and deletions, which is captured in the SNP mapping in Figure 3b.
2. For each patient, we recoded their mutation status into one of the tokens from the dictionary.
3. For each SNP, we gather the tokens for all the patients into a vector of tokens and use it to learn embeddings.

### 2.4 Model parameters

We train the model using CLIP architecture as shown in Algorithm 1. We extract feature representations of each modality using the SNP and IDP encoders respectively, as discussed above, for each batch of the data. We obtain 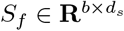 and 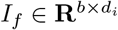 where there are *b* pairs in each batch and *d*_*s*_, *d*_*i*_ are sizes of the tokenized embeddings. We normalize the embeddings after learning joint multimodal embeddings using the projection weight matrices 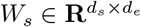 and 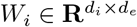

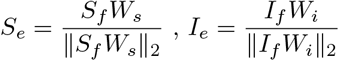

We then computed the cosine similarities between *S*_*e*_ and *I*_*e*_. Thereafter, we computed a symmetric loss function by computing using two cross-entropy losses, one for each modality and averaging them. The ground truth is established as the index of predictions of the model, i.e. it is expected that the model will predict the corresponding IDP position given a SNP position. We used the Adam optimizer with learning rate warm up and weight decay modifiers.. Hyperparameter tuning was used to further improve the pair learning capabilities from `COMICAL`. The library Ray Tune was used to perform efficient hyperparameter tuning. The best hyperparameters can be found in Table 1. We observed similar hyperparameters across thresholds of SNPs with the major differences being in the training hyperparameters, namely learning rate, warm up steps and weight decay.

**Table 1:** Best hyperparameters obtained through Ray Tune for the top 0.5% and top 2% SNPs.

|  | Top 0.5% SNPs | Top 2% SNPs |
| --- | --- | --- |
| **Batch size** | 4096 | 4096 |
| **Model dimensions** | 64 | 64 |
| **Number of Transformer layers** | 2 | 2 |
| **Attention heads per layer** | 4 | 4 |
| **Feed-forward dimensions** | 64 | 64 |
| **Learning rate** | 0.0033 | 0.0001 |
| **Warmup steps** | 236 | 343 |
| **Weight decay** | 0.0015 | 0.0183 |

### 2.5 Data

We used the UKB dataset [26] to obtain 154 IDPs derived from the T1 structural brain MRIs spanning 47,420 samples. Of these samples, we performed genomic quality control such as removing samples with missing data (more than 10%), high variance of heterozygosity rate, closely related individuals, etc. to finally obtain 40,426 samples of European ancestry. We performed principal component analysis (PCA) from the genomic data of pruned markers using `TeraPCA` [6] to obtain forty PCs for using as covariates in downstream tasks in the model. We then selected 5,632 variants previously associated with the eight neurological diseases from the GWAS catalog and checked for missingness in SNPs, low minor allele frequency (< 0.05), Hardy-Weinberg equilibrium (*p* < 1×10^−16^), etc. to finally obtain 5,603 variants for the pre-training phase. We selected ten neurological diseases across the 40,426 samples and used eight of them in the pair-making process. The number of cases for each neurological disease is given in Table 2.

**Table 2:** Number of cases for each neurological diseases and their respective percentage in the dataset of 40,426 samples.

| Disease | AD | ADHD | ASD | BD | CBVD | MD | MS | PD | Stroke | SZ | UD |
| --- | --- | --- | --- | --- | --- | --- | --- | --- | --- | --- | --- |
| Count | 12 | 17 | 7 | 127 | 489 | 10 | 141 | 85 | 489 | 55 | 4107 |
| Percent | 0.0003 | 0.0004 | 0.0002 | 0.0031 | 0.0127 | 0.0003 | 0.0034 | 0.00021 | 0.0121 | 0.0014 | 0.1016 |

### 2.6 Post processing

To compute *p*-values, we captured frequencies per batch of correct SNP-IDP pairs that `COMICAL` identified during evaluation of the test set. After collating all the frequencies per pair, we performed a one-way *χ*^2^ test. Due to substantial differences in frequencies among important pairs and less important pairs, we corrected the *χ*^2^ values by dividing by the number of observations and then computed adjusted *p*-values. The SNP-IDP pairs, frequencies and *p*-values are included in the Supplementary Table 1.

### 2.7 Prediction tasks

We sought to use `COMICAL` learned transformer embeddings for both the modalities of SNPs and IDPs and predict clinical outcomes as well as compute risk score estimates.

#### Clinical outcomes

We simply concatenated the transformer encoded embeddings from `COMICAL` and used it as an input to predict clinical outcome. In this case, we used these embeddings to predict eleven outcomes: eight neurological diseases we considered in the pair-making process; two additional unseen outcomes (stroke and schizophrenia), and another meta-disease outcome which is a combination of all the diseases by performing a logical AND operation on the indicator variables of the ten outcomes.

As specific encoded representations for SNPs and IDPs are learned using `COMICAL`, we concatenated them as input to a neural network with a single hidden layer, and a final layer to output the logits. The SNP and IDP encoders were frozen, leading to a lightweight training of just the classifier network. We used age, sex, and the top forty PCs as covariates in this analysis. The framework of outcome prediction is shown in Figure 4. We use a cross entropy loss between the true disease status and the predicted outcome.

**Fig. 4:**
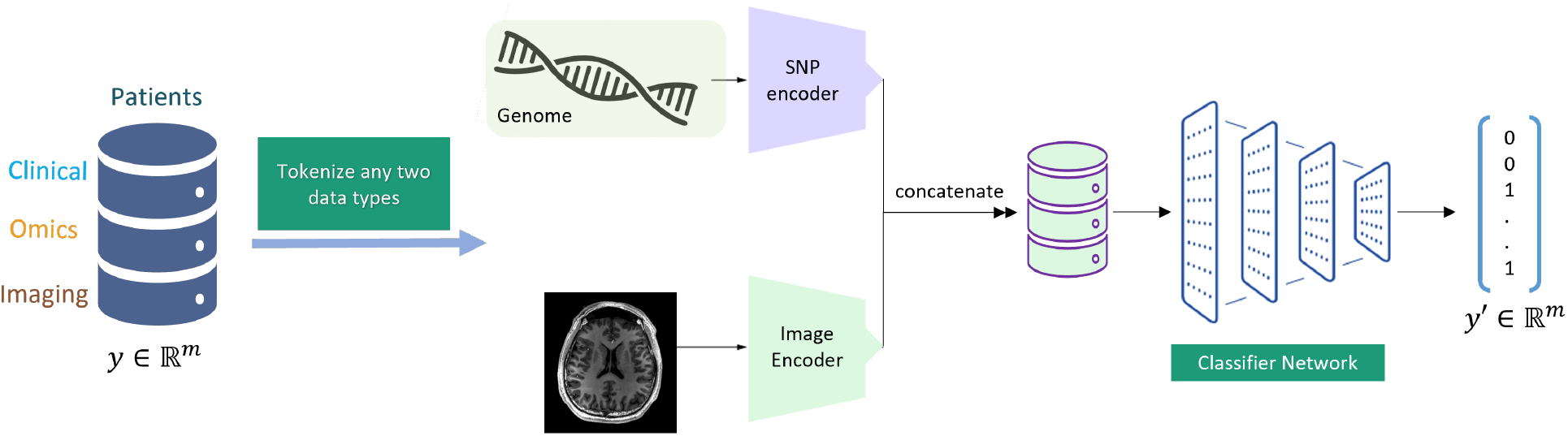
Overview of outcome prediction mechanism in `COMICAL`. We concatenate the encodings from both modalities to predict clinical outcome.

#### Risk scores

We performed risk scoring for 5 diseases of interest, namely AD, BD, MS, PD, SZ. Using the concatenated embeddings from the SNPs and IDPs from `COMICAL` we built multi-omics representations for each individual in the dataset. Then using a neural network with a single hidden layer, we trained the model to regress the enhanced PRS (ePRS) set available from the UK Biobank [28] for the 5 diseases. We used 10% of the top SNPs from GWAS catalog to train `COMICAL` and used 1280 samples for training, 320 for validation and 4992 testing. The training set was defined as samples with a diagnosis of a disorder and an equal amount of healthy controls. The test set was comprised of the remaining healthy control samples; thus ensuring completely unseen data for evaluation. Mean squared error (MSE) was used as loss to be minimized, and *R*^2^ was used as the goodness-of-fit metric. The risk scoring process is outlined in Algorithm 2. We evaluated the performance of the `COMICAL` risk score by fitting a least square regression with the ePRS using `statsmodels` [23] and included covariates of age, sex, smoking status, BMI, height, and the top forty PCs.

##### Algorithm 2

`COMICAL`: risk score estimation

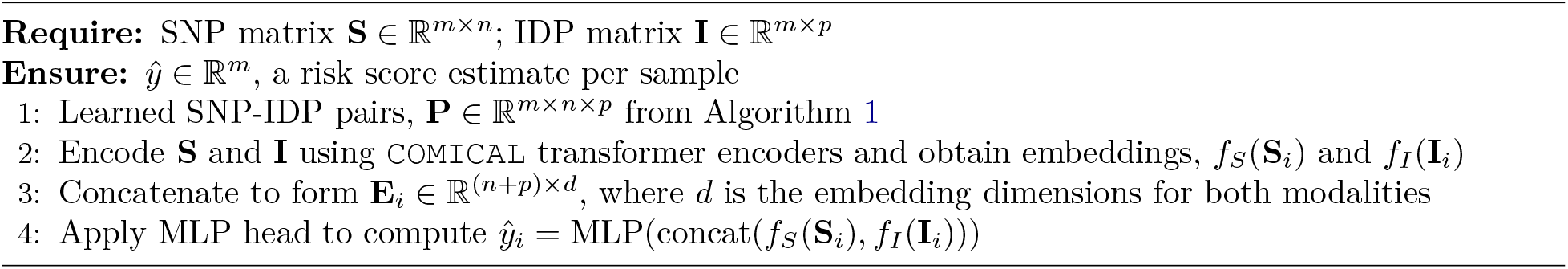

## 3 Results

### 3.1 Learning pairs in UK Biobank

In our experiments utilizing data from the UK Biobank, our objective was to evaluate `COMICAL`’s performance, focusing on its ability to learn associations. We varied the amount of data that `COMICAL` would ingest and learn from only including a certain percentage of the top SNPs according to the strength of association from the GWAS catalog. We created pairs using the process outlined above and processed them using Algorithm 1. As an example, utilizing only the top 0.5 percent of SNPs resulted in the generation of over 15 million pairs. After training the model, we evaluated how well `COMICAL` learned the SNP-IDP pairs using the test set. `COMICAL` correctly identified the SNP-IDP pairs with 97.3% accurately on the test set when trained using the top 0.5 percent SNPs. After hyperparameter tuning and training with the optimal parameters (Table 1), we observed `COMICAL` was able to predict every SNP-IDP pair in the test set, with a 100% accuracy. We also measured the execution time for each experiment using `COMICAL` including the pair-making between IDPs and SNPs. As the number of associations increases, there is a substantial growth in the quantity of SNP-IDP pairs which increases the amount of information for the model to learn from, and this is duly reflected in the overall execution time. Training time was directly proportional to the number of A100 GPUs that were used in training. For 5% top SNPs in the training set, `COMICAL` took approximately 85 minutes to train with 3 A100 GPUs. However, for a single GPU, it took 218 minutes.

### 3.2 Comparison with GWAS summary statistics

We compared the top associations with respect to *p*-values with GWAS summary statistics from ENIGMA consortium, specifically, ENIGMA2 [13] and ENIGMA3 [11]. We found the SNP rs1516725, in the region of DGKG, which is associated with obesity in GWAS catalog, being mapped to the volume of grey matter in right ventral striatum (*p <* 10^−26^). This SNP was found to be associated with the mean volume of hippocampus in ENIGMA2 summary statistics (*p* < 9 *×* 10^−4^). Right ventral striatum is involved in reward processing and motivation in the brain, whereas hippocampus deals with contextual memories, which can include memories of reward and reinforcement. Hippocampus often works together with the ventral striatum and both are related to obesity [3, 9]. The reward and motivation impulse in the brain is often linked with affinity towards weight gain and increased obesity risk [9]. Another such association is rs266058 which is associated with the volume of grey matter in left amygdala (*p* < 2×10^−24^) and strongly associated with ADHD in GWAS catalog. It is known that ADHD patients tend to have smaller amygdala volumes compared to healthy controls [27]. This SNP was associated with the isthmus cingulate cortex in ENIGMA3 summary statistics (*p* < 2×10^−4^). A large brain imaging study of the cortex in ADHD called ENIGMA-ADHD [14] suggested to study the network of orbitofrontal cortex, cingulate, and amygdala for understanding deficient emotional self-regulation in ADHD patients [24]. `COMICAL` was able to find this subtle connection which had not been found earlier in ADHD GWAS studies. Similar to this, `COMICAL` was able to find many other connections such as rs993137, another SNP strongly associated with ADHD was paired with volume of grey matter in left amygdala in `COMICAL` (*p* < 3×10^−11^) and mean volume of inferior temporal cortex in ENIGMA2. The inferior temporal cortex is strongly associated with behavioral inhibition and hence connected to ADHD [1]. The full results of the significant learned pairs from `COMICAL` and their mapping to IDPs obtained from ENIGMA studies is shared in Supplementary Table 2. We observed that on an average, across different GWAS catalog thresholds, 50% of significant SNPs obtained from `COMICAL` were found to have associations in ENIGMA summary statistics (*p* < 5 *×* 10^−4^).

### 3.3 Cross-disorder prediction

We used `COMICAL` to leverage the learned embeddings of the SNP-IDP pairs and used them to predict disease outcome. The key knowledge of the SNP-IDP relationships encoded in the embeddings was beneficial to the predictive capabilities of the data. Using the SNP and IDP encoders from `COMICAL` we were able to train a small multi-layer perceptron (MLP) to predict disease outcome states and compared it with a baseline of using the original SNPs and IDPs without the encoding and evaluate it in the same MLP architecture. As seen in Figure 5, using the `COMICAL` trained encoders with an MLP achieves very high performances when compared to the baseline for ADHD, ASD, MD, and SZ. It is exciting to see the high performance of SZ compared to the baseline, as SZ is an unseen disease for `COMICAL`. This shows the promising capabilities of `COMICAL` to implicitly learn disease SNP-IDP relationships that can be used for unseen outcomes of interest. Additionally, we see comparable performances on AD, BPD, UD, MS, Stroke and meta. Finally, we see a lower performance on PD when compared to the baseline. The low performances of `COMICAL` and baselines could be in part explained due to low number of cases in the dataset (Table 2), as UK Biobank is a disease agnostic dataset. The relative performance with respect to the baseline was captured using net reclassification index (NRI) as the difference between the area under the curve value obtained by `COMICAL` vs. the baseline (Figure 6). Our current results, show the promising capabilities of `COMICAL` towards downstream tasks, as a potential foundation model in the SNP-IDP domain.

**Fig. 5:**
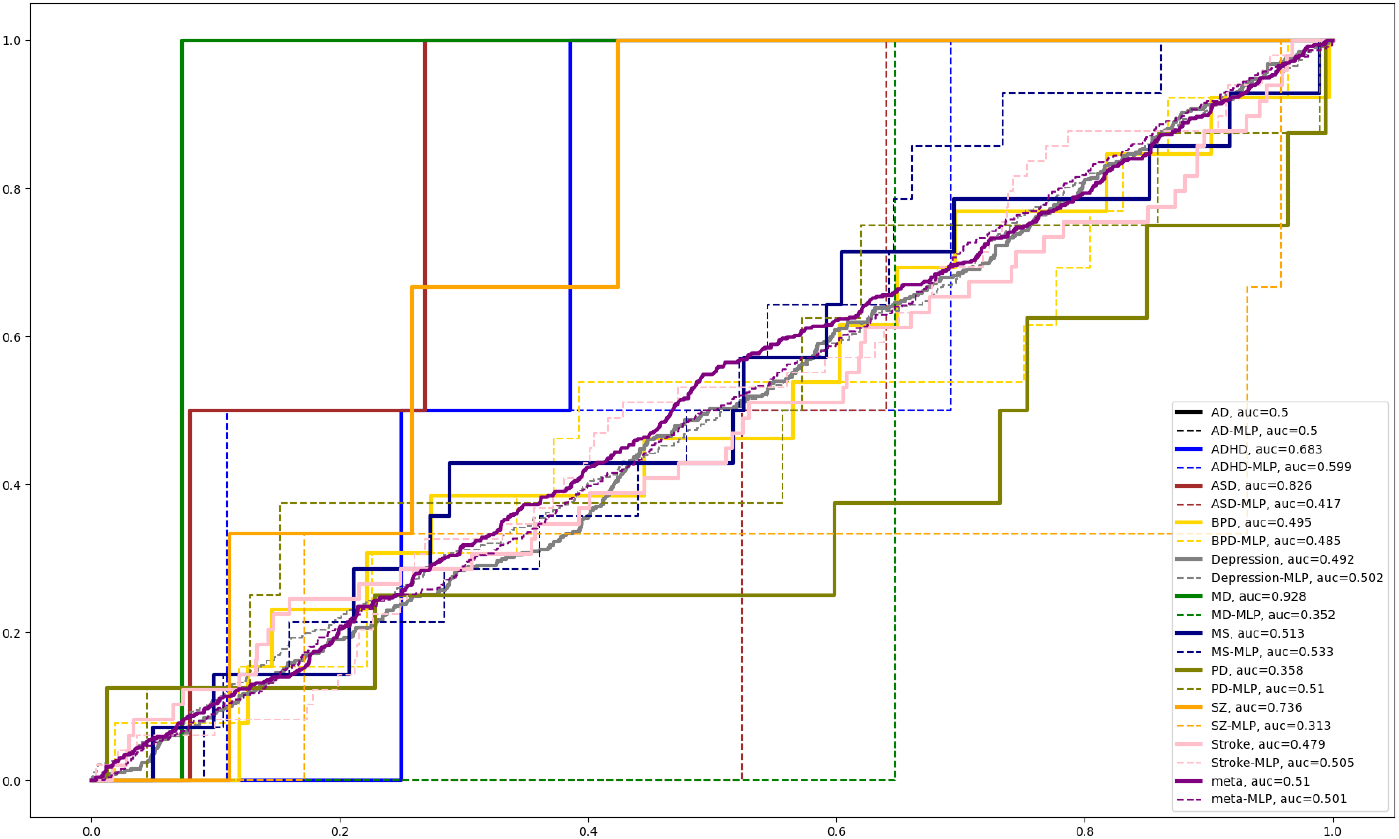
Receiver operating characteristic curve plots for each one of the diseases using MLP plus `COMICAL` encoders, captured by solid lines and baseline MLPs (denoted as disease ‘-’ MLP), captured by dashed lines for comparison.

**Fig. 6:**
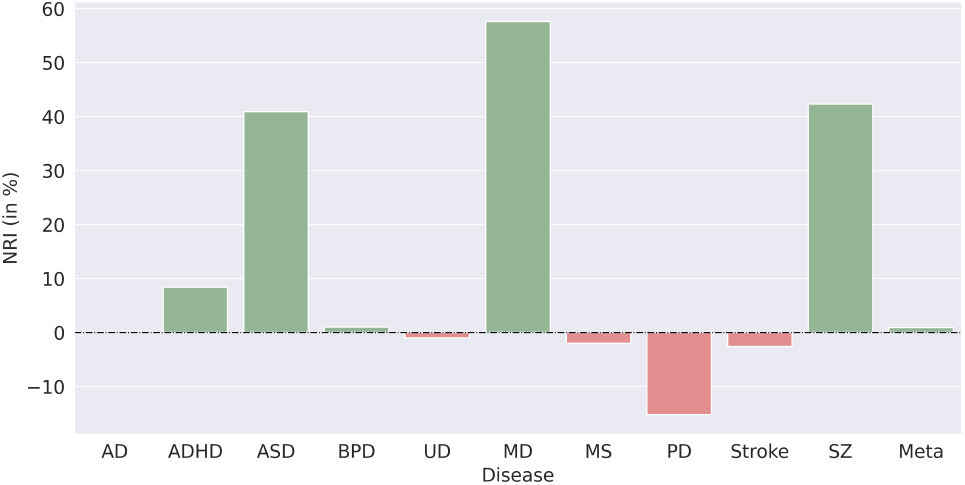
Net reclassification index (NRI) of AUC in `COMICAL` vs. the baseline across the eleven clinical outcomes.

Similarly, `COMICAL` showed promising capabilities at predicting risk scoring for 5 different neurological disorders. More importantly, it was able to be tuned on predicting for one disorder and then effectively estimate risk scores on a different one, as shown in Table 3. The similar *R*^2^ across disorders suggest that as there are several shared patterns across neurological conditions it is feasible to generalize across disorders. Additionally, the results suggested that the target disorder is highly influential in the abilities from `COMICAL` to perform accurate risk scoring. For example, SZ consistently achieves the highest *R*^2^, while also achieving high AUCs as seen in Figure 5.

**Table 3:** *R*^2^ values for risk score prediction task are shown. When training COMICAL on the prediction task of risk scoring for a given disease, COMICAL can then be used to predict the risk score on a different disease. Disease used for training is show on the rows while the adapted disease is shown as columns. Significance at *p* < 0.05.

|  | AD | BD | MS | PD | SZ |
| --- | --- | --- | --- | --- | --- |
| AD | 0.696 | 0.687 | 0.666 | 0.698 | 0.709 |
| BD | 0.693 | 0.691 | 0.662 | 0.698 | 0.714 |
| MS | 0.693 | 0.689 | 0.664 | 0.698 | 0.713 |
| PD | 0.697 | 0.663 | 0.697 | 0.707 | 0.708 |
| SZ | 0.694 | 0.691 | 0.659 | 0.699 | 0.708 |

## 4 Discussion

We have developed a multi-modal foundation model for neurological disorders with a contrastive learning approach leveraging multi-omics data to generate many-to-many associations between genetic markers and brain imaging phenotypes. Our approach discovered several associations between SNPs and IDPs that are validated in the literature to have strong associations with neurological and psychiatric conditions. This demonstrates the capacity of `COMICAL` to unravel intricate interactions within complex disorders which are often missed by traditional association studies such as GWAS, eQTL, etc. `COMICAL` was able to find subtle connections between SNPs and IDPs which were implicitly mentioned in previous studies [9, 14, 24], as well as recover well known associations between them. In the cross-disorder prediction task, `COMICAL` either performed upto 60% better than the baseline or performed almost as well, except the exception of PD. On top of this, `COMICAL` was able to perform very well (73.6%) on predicting an unseen disease, SZ from the learned pairs, highlighting the discriminative aspects of foundation models. Some of the disease outcomes had very low representation in the dataset (Table 2), which might play a role in reduced performance, in some cases. `COMICAL` showed very promising results when predicting risk scores for five neurological disorders for which ePRS data was available. It was able to predict across disorders when trained only on one of them (Table 3). This elucidates the path towards foundation model capabilities in multi-omics analysis. `COMICAL` facilitates transfer learning across modalities as well as in unseen, held-out data with the pre-trained models. `COMICAL` trained on UKB data can be used to predict disorders on neurological cohorts such as Alzheimer’s Disease Neuroimaging Initiative [19], Parkinson’s Progression Markers Initiative [16], ENIGMA [29], etc. and enable biomarker discovery.

Despite the novelty and innovation that our approach provides, it is not without limitations. One shortcoming is a result of the pair-making process. Depending on the modalities that are being analyzed, the embedding space becomes incredibly large when generating possible pairs. This led to our design choice to limit the model to learn using the top SNPs when evaluating on the UK Biobank data. We note that this has resulted in sub-optimal performance in some cases and we expect to engineer architectural advances in `COMICAL` to train it with even larger space of parameters while ingesting whole genome sequencing data. Another direction for future research would be applying `COMICAL` to other large biobanks with neurological diseases or perform pre-training on other complex diseases such as cancer, cardiovascular disease, etc. This would further demonstrate the versatility `COMICAL` has and illustrate its significance to the domain of multimodal foundation models. We also expect to improve the architecture of `COMICAL` by trying other encoders and tokenizers which preserve domain information in multimodal foundation models. Although we showcase `COMICAL` using genomic and imaging markers, we can extend its capabilities to learn associations between genomic markers and proteins, genes and methylation information, etc. and thus `COMICAL` is domain agnostic.

## 5 Conclusion

Multi-omics data analysis is essential towards understanding complex diseases such as neurological disorders with their high comorbidity. However, discovering the intricate relationships between genomic markers and brain imaging phenotypes can be very challenging as there are large sets of underlying mechanisms. Commonly used methods such as GWAS are often single-marker association studies, not observing multi-way or many-to-many associations. `COMICAL`, a foundation model tuned to identify associations of SNPs and IDPs in large cohorts allows us to learn the complex many-to-many genotype-phenotype relationships and use these towards downstream tasks. Our results show the strong capabilities from `COMICAL` to learn associations between SNPs and IDPs in a self-supervised manner using UK Biobank data, and its promising capabilities towards disease outcome prediction and estimating a novel multi-modal, multi-omics risk score. `COMICAL` enables transfer learning and accelerates discovery of therapeutic targets with the availability of a pre-trained model.

## Data Availability

Data is available from UK Biobank.

## 6. Code and Data availability

Data is available upon request from UK Biobank. Code is available in https://github.com/IBM/comical.

## Acknowledgements

D.M.R performed this work as part of a summer internship at IBM Research.

## Author contributions

A.B conceived this project. D.M.R implemented the training and evaluation phases of `COMICAL`. M.B performed the pair making task. A.B, D.M.R, M.B wrote the manuscript. All authors participated in discussions and reviewed the manuscript.

