## Supplemental Tables for "A Multimodal Foundation Model for Discovering Genetic Associations with Brain Imaging Phenotypes"

| SNPs | IDPs (Vol. of grey matter) | adj-p |
|---|---|---|
| rs2568958 | Planum_Temporale_(right) | 8.28E-48 |
| **rs9919558** | Supramarginal_Gyrus_(right) | 7.32E-41 |
| **rs266058** | Amygdala_(left) | 2.50E-24 |
| **rs3104373** | Middle_Temporal_Gyrus,_temporooccipital_part_(left) | 3.62E-27 |
| **rs2232423** | Putamen_(right) | 6.28E-10 |
| **rs1516725** | Ventral_Striatum_(right) | 2.93E-26 |
| **rs9919558** | Superior_Temporal_Gyrus_(right) | 5.50E-18 |
| **rs9919558** | Middle_Temporal_Gyrus_(left) | 2.07E-15 |
| **rs2568958** | Supramarginal_Gyrus_(left) | 0.005939 |
| **rs429358** | Supracalcarine_Cortex_(right) | 0.015023 |
| **rs356182** | Frontal_Medial_Cortex_(left) | 4.35E-47 |
| **rs191800971** | Brain-Stem | 2.18E-14 |
| **rs993137** | Amygdala_(left) | 3.25E-11 |
| **rs9919558** | Heschl's_Gyrus_(includes_H1_H2)_(left) | 0.001256 |
| **rs10119** | Supracalcarine_Cortex_(right) | 3.21E-05 |
| **rs1516725** | Superior_Temporal_Gyrus_(left) | 0.001082 |
| **rs1516725** | Supramarginal_Gyrus_(left) | 0.03066 |
| **rs3104373** | Precentral_Gyrus_(right) | 0.005137 |
| **rs2075650** | Supracalcarine_Cortex_(right) | 0.000187 |
| **rs2568958** | Temporal_Pole_(right) | 0.001371 |
| **rs2568958** | Occipital_Fusiform_Gyrus_(left) | 0.044203 |
| **rs2568958** | VIIIb_Cerebellum_(right) | 0.008824 |
| **rs30266** | Brain-Stem | 0.000473 |
| **rs2568958** | Thalamus_(right) | 2.00E-30 |
| **rs9919558** | Amygdala_(left) | 0.017519 |
| **rs2568958** | VI_Cerebellum_(left) | 1.34E-45 |
| **rs2568958** | Temporal_Occipital_Fusiform_Cortex_(right) | 4.77E-24 |
| **rs2092563** | Brain-Stem | 3.12E-28 |
| **rs2568958** | Supracalcarine_Cortex_(left) | 4.19E-51 |
| **rs9919558** | VI_Cerebellum_(right) | 1.02E-47 |
| **rs9919558** | Pallidum_(left) | 0.004954 |
| **rs3104373** | Juxtapositional_Lobule_Cortex_(right) | 4.73E-21 |
| **rs1516725** | Supramarginal_Gyrus_(right) | 2.03E-21 |
| **rs9919558** | Insular_Cortex_(left) | 4.63E-20 |
| **rs814573** | Supracalcarine_Cortex_(right) | 0.000306 |
| **rs9919558** | Parietal_Operculum_Cortex_(left) | 2.25E-28 |
| **rs3104373** | X_Cerebellum_(left) | 0.032799 |
| **rs1516725** | V_Cerebellum_(right) | 3.77E-23 |
| **rs356182** | VIIIa_Cerebellum_(left) | 3.24E-45 |
| **rs9919558** | Frontal_Operculum_Cortex_(right) | 1.45E-23 |
| **rs356182** | Thalamus_(left) | 1.27E-23 |
| **rs356182** | V_Cerebellum_(left) | 1.59E-23 |

Supplementary Table 1: Significant SNP-IDP pairs obtained from COMICAL

| SNPs | IDPs (Vol. of grey matter) | adj-p |
|---|---|---|
| rs993137 | Amygdala_(left) | 1.73E-54 |
| rs1081105 | Supracalcarine_Cortex_(right) | 4.26E-70 |
| rs9919558 | Lingual_Gyrus_(right) | 2.85E-24 |
| rs2568958 | X_Cerebellum_(vermis) | 2.03E-24 |
| rs28606370 | Amygdala_(left) | 1.17E-19 |
| rs3129889 | Juxtapositional_Lobule_Cortex_(right) | 1.04E-58 |
| rs9919558 | Supramarginal_Gyrus_(right) | 6.84E-24 |
| rs2568958 | Caudate_(right) | 0.003473 |
| rs61902811 | Brain-Stem | 5.88E-28 |
| rs9919558 | Amygdala_(left) | 9.09E-34 |
| rs9919558 | Parietal_Operculum_Cortex_(left) | 1.35E-29 |
| rs2568958 | Inferior_Temporal_Gyrus,_temporooccipital_part | 9.55E-18 |
| rs1516725 | V_Cerebellum_(right) | 1.93E-64 |
| rs1004787 | Amygdala_(left) | 6.21E-32 |
| rs30266 | Brain-Stem | 0.008338 |
| rs28399637 | Supracalcarine_Cortex_(right) | 2.77E-26 |
| rs75627662 | Supracalcarine_Cortex_(right) | 2.75E-34 |
| rs2568958 | Planum_Polare_(right) | 2.35E-30 |
| rs157580 | Supracalcarine_Cortex_(right) | 6.66E-07 |
| rs3104373 | Precentral_Gyrus_(right) | 0.000631 |
| rs2568958 | Cuneal_Cortex_(left) | 8.83E-26 |
| rs356182 | Thalamus_(left) | 1.61E-09 |
| rs3104373 | Middle_Temporal_Gyrus,_temporooccipital_part | 0.011644 |
| rs2075650 | Supracalcarine_Cortex_(right) | 6.18E-11 |
| rs356182 | Juxtapositional_Lobule_Cortex_(left) | 0.010394 |
| rs10119 | Supracalcarine_Cortex_(right) | 0.006895 |
| rs2568961 | Putamen_(right) | 0.005445 |
| rs1568452 | Brain-Stem | 0.001911 |
| rs9919558 | Superior_Temporal_Gyrus_(right) | 0.000926 |
| rs356182 | Frontal_Medial_Cortex_(left) | 4.92E-12 |
| rs2568958 | Inferior_Frontal_Gyrus,_pars_opercularis_(left) | 0.002218 |
| rs2568958 | VIIIb_Cerebellum_(right) | 0.013896 |
| rs1516725 | Superior_Temporal_Gyrus_(left) | 0.016962 |
| rs2568958 | Crus_II_Cerebellum_(vermis) | 0.012634 |
| rs191800971 | Brain-Stem | 0.01202 |
| rs3888190 | Superior_Temporal_Gyrus_(left) | 1.14E-06 |
| rs1516725 | Ventral_Striatum_(right) | 0.004642 |
| rs2568958 | Crus_II_Cerebellum_(left) | 0.008324 |
| rs1516725 | Supramarginal_Gyrus_(left) | 0.01498 |
| rs9271366 | Juxtapositional_Lobule_Cortex_(right) | 0.02588 |
| rs356182 | Inferior_Temporal_Gyrus_(right) | 0.013831 |
| rs2568958 | Crus_I_Cerebellum_(left) | 0.011502 |
| rs6452785 | Amygdala_(left) | 0.000355 |
| rs3104373 | Juxtapositional_Lobule_Cortex_(right) | 3.13E-26 |
| rs1095626 | Brain-Stem | 3.14E-25 |
| rs356182 | Pallidum_(right) | 1.36E-24 |

| | | |
|---|---|---|
| rs266058 | Amygdala_(left) | 0.000835 |
| rs2568958 | VIIb_Cerebellum_(vermis) | 0.002052 |
| rs2568958 | Occipital_Fusiform_Gyrus_(left) | 1.59E-52 |
| rs1516725 | Supramarginal_Gyrus_(right) | 8.15E-64 |
| rs2568958 | Parahippocampal_Gyrus_(right) | 1.36E-60 |
| rs2568958 | Supracalcarine_Cortex_(left) | 1.50E-53 |
| rs356182 | V_Cerebellum_(left) | 1.57E-33 |
| rs2568958 | Inferior_Temporal_Gyrus_(left) | 1.74E-60 |
| rs2232423 | Putamen_(right) | 2.50E-28 |
| rs41289512 | Supracalcarine_Cortex_(right) | 2.91E-24 |
| rs2092563 | Brain-Stem | 8.16E-20 |
| rs356182 | VIIIa_Cerebellum_(left) | 1.53E-26 |
| rs2568958 | Brain-Stem | 1.52E-27 |
| rs356203 | Frontal_Medial_Cortex_(left) | 6.33E-29 |
| rs2568958 | Hippocampus_(right) | 8.12E-20 |
| rs814573 | Supracalcarine_Cortex_(right) | 3.97E-15 |
| rs9919558 | VI_Cerebellum_(right) | 4.49E-28 |
| rs2568958 | Middle_Frontal_Gyrus_(right) | 6.41E-27 |
| rs429358 | Supracalcarine_Cortex_(right) | 4.95E-14 |
| rs3104373 | X_Cerebellum_(left) | 2.09E-25 |
| rs9919558 | Heschl's_Gyrus_(includes_H1_H2)_(left) | 7.54E-13 |
| rs2568958 | Inferior_Frontal_Gyrus,_pars_opercularis_(right) | 1.45E-11 |
| rs9919558 | Amygdala_(right) | 7.38E-05 |
| rs557042 | Putamen_(right) | 0.017487 |

| SNPs | IDPs | adj-p |
|---|---|---|
| rs1516725 | V_Cerebellum_(right) | 9.18E-27 |
| rs356182 | Middle_Temporal_Gyrus_(left) | 2.30E-27 |
| rs356182 | Pallidum_(right) | 0.0185 |
| rs406456 | Supracalcarine_Cortex_(right) | 8.41E-35 |
| rs3104373 | Precentral_Gyrus_(right) | 7.56E-40 |
| rs1516725 | Ventral_Striatum_(right) | 1.65E-38 |
| rs7111031 | Brain-Stem | 0.000777 |
| rs356182 | Thalamus_(left) | 8.52E-30 |
| rs951740 | Amygdala_(left) | 1.46E-22 |
| rs3104373 | Middle_Temporal_Gyrus,_temporooccipital_part_(left) | 6.77E-39 |
| rs34637584 | Frontal_Medial_Cortex_(left) | 4.61E-37 |
| rs3888190 | Superior_Temporal_Gyrus_(left) | 2.84E-75 |
| rs3129889 | Juxtapositional_Lobule_Cortex_(right) | 6.36E-23 |
| rs1081105 | Supracalcarine_Cortex_(right) | 2.47E-39 |
| rs1065853 | Supracalcarine_Cortex_(right) | 0.003438 |
| rs1381287 | Amygdala_(left) | 0.02698 |
| rs9919558 | Superior_Temporal_Gyrus_(right) | 0.00072 |
| rs28399637 | Supracalcarine_Cortex_(right) | 0.024091 |
| rs2568958 | Middle_Frontal_Gyrus_(right) | 0.01778 |
| rs3925681 | Supracalcarine_Cortex_(right) | 0.011131 |
| rs2568958 | Inferior_Frontal_Gyrus,_pars_opercularis_(left) | 2.57E-10 |
| rs266058 | Amygdala_(left) | 0.021136 |
| rs12907546 | Amygdala_(left) | 0.00148 |
| rs9919558 | Supramarginal_Gyrus_(right) | 0.021795 |
| rs11688767 | Brain-Stem | 0.018528 |
| rs6859 | Supracalcarine_Cortex_(right) | 0.014111 |
| rs9919558 | Parietal_Operculum_Cortex_(left) | 4.93E-05 |
| rs1800693 | Juxtapositional_Lobule_Cortex_(right) | 0.014212 |
| rs2568958 | Insular_Cortex_(right) | 0.001382 |
| rs9919558 | Heschl's_Gyrus_(includes_H1_H2)_(left) | 0.014005 |
| rs1516725 | Superior_Temporal_Gyrus_(left) | 0.019442 |
| rs2568958 | Temporal_Occipital_Fusiform_Cortex_(left) | 0.010905 |
| rs10801908 | Juxtapositional_Lobule_Cortex_(right) | 0.008381 |
| rs438613 | Juxtapositional_Lobule_Cortex__(right) | 0.00313 |
| rs2568958 | IX_Cerebellum_(vermis) | 0.006697 |
| rs2568958 | Temporal_Fusiform_Cortex_(right) | 1.03E-05 |
| rs1516725 | Supramarginal_Gyrus_(left) | 1.37E-31 |
| rs458806 | Amygdala_(left) | 0.016444 |
| rs75627662 | Supracalcarine_Cortex_(right) | 0.021718 |
| rs9919558 | VI_Cerebellum_(right) | 0.006418 |
| rs7259620 | Supracalcarine_Cortex_(right) | 0.001967 |
| rs2568961 | Putamen_(right) | 0.000191 |
| rs4420638 | Supracalcarine_Cortex_(right) | 9.20E-08 |
| rs3104373 | Juxtapositional_Lobule_Cortex_(right) | 0.006806 |
| rs157580 | Supracalcarine_Cortex_(right) | 0.017109 |
| rs9271366 | Juxtapositional_Lobule_Cortex_(right) | 0.004584 |

| | | |
|---|---|---|
| rs41289512 | Supracalcarine_Cortex_(right) | 0.009251 |
| rs2075650 | Supracalcarine_Cortex_(right) | 9.71E-29 |
| rs356182 | VIIIa_Cerebellum_(left) | 7.21E-74 |
| rs2568958 | Parietal_Operculum_Cortex_(right) | 0.000415 |
| rs10890020 | Brain-Stem | 0.001554 |
| rs2927468 | Supracalcarine_Cortex_(right) | 0.000429 |
| rs2232423 | Putamen_(right) | 8.37E-36 |
| rs9834970 | Superior_Temporal_Gyrus_(left) | 6.48E-39 |
| rs557042 | Putamen_(right) | 2.94E-05 |
| rs3104373 | X_Cerebellum_(left) | 2.19E-35 |
| rs2568958 | Cuneal_Cortex_(left) | 0.00067 |
| rs2568958 | Middle_Frontal_Gyrus_(left) | 0.00014 |
| rs200965 | Putamen_(right) | 0.00013 |
| rs9919558 | Amygdala_(left) | 5.33E-06 |
| rs356182 | V_Cerebellum_(left) | 3.85E-29 |
| rs2092563 | Brain-Stem | 3.97E-43 |
| rs1516725 | Supramarginal_Gyrus_(right) | 2.61E-56 |
| rs9919558 | Middle_Temporal_Gyrus_(left) | 8.58E-57 |
| rs429358 | Supracalcarine_Cortex_(right) | 1.78E-06 |
| rs9919558 | Pallidum_(left) | 9.38E-32 |
| rs356182 | Inferior_Temporal_Gyrus_(right) | 0.009019 |
| rs356182 | Frontal_Medial_Cortex_(left) | 7.50E-26 |
| rs62401383 | Putamen_(right) | 4.19E-56 |
| rs34311866 | Frontal_Medial_Cortex_(left) | 2.04E-57 |
| rs356182 | Juxtapositional_Lobule_Cortex_(left) | 7.87E-05 |
| rs4856605 | Amygdala_(left) | 0.018259 |
| rs814573 | Supracalcarine_Cortex_(right) | 1.18E-20 |
| rs200949 | Brain-Stem | 4.83E-63 |
| rs1004787 | Amygdala_(left) | 7.24E-06 |

| SNPs | IDPs (Vol. of grey matter) | adj-pvalue | POS | Enigma IDP |
|---|---|---|---|---|
| rs1516725 | Ventral_Striatum_(right) | 2.93E-26 | 3:185824004 | MeanHippocampus |
| rs266058 | Amygdala_(left) | 2.50E-24 | 2:104086428 | Mean_isthmuscingulate_surfavg |
| rs266058 | Amygdala_(left) | 2.50E-24 | 2:104086428 | Mean_Full_Thickness |
| rs1516725 | V_Cerebellum_(right) | 3.77E-23 | 3:185824004 | MeanHippocampus |
| rs1516725 | Supramarginal_Gyrus,_posterior_division_(right) | 2.03E-21 | 3:185824004 | MeanHippocampus |
| rs993137 | Amygdala_(left) | 3.25E-11 | 3:85499035 | Mean_inferiortemporal_surfavg |
| rs993137 | Amygdala_(left) | 3.25E-11 | 3:85499035 | Mean_Full_Thickness |
| rs10119 | Supracalcarine_Cortex_(right) | 3.21E-05 | 19:45406673 | MeanAccumbens |
| rs2075650 | Supracalcarine_Cortex_(right) | 0.000186506 | 19:45395619 | MeanAccumbens |
| rs2075650 | Supracalcarine_Cortex_(right) | 0.000186506 | 19:45395619 | MeanHippocampus |
| rs1516725 | Superior_Temporal_Gyrus,_anterior_division_(left) | 0.001082216 | 3:185824004 | MeanHippocampus |
| rs429358 | Supracalcarine_Cortex_(right) | 0.015022532 | 19:45411941 | MeanAmygdala |
| rs429358 | Supracalcarine_Cortex_(right) | 0.015022532 | 19:45411941 | MeanAccumbens |
| rs429358 | Supracalcarine_Cortex_(right) | 0.015022532 | 19:45411941 | MeanHippocampus |
| rs429358 | Supracalcarine_Cortex_(right) | 0.015022532 | 19:45411941 | MeanThalamus |
| rs1516725 | Supramarginal_Gyrus,_posterior_division_(left) | 0.030660033 | 3:185824004 | MeanHippocampus |

Supplementary Table 2: ENIGMA mapped SNP-IDP pairs for 0.5% top SNPs in GWAS catalog

| SNPs | IDPs (Vol. of grey matter) | adj-pvalue | MARKER | Enigma IDF |
|---|---|---|---|---|
| rs3888190 | Superior_Temporal_Gyrus_(left) | 2.84E-75 | 16:28889486 | MeanCaud |
| rs3888190 | Superior_Temporal_Gyrus_(left) | 2.84E-75 | 16:28889486 | MeanPutar |
| rs3888190 | Superior_Temporal_Gyrus_(left) | 2.84E-75 | 16:28889486 | MeanAccur |
| rs3888190 | Superior_Temporal_Gyrus_(left) | 2.84E-75 | 16:28889486 | Mean_infe |
| rs34311866 | Frontal_Medial_Cortex_(left) | 2.04E-57 | 4:951947 | Mean_late |
| rs1516725 | Supramarginal_Gyrus_(right) | 2.61E-56 | 3:185824004 | MeanHippo |
| rs1081105 | Supracalcarine_Cortex_(right) | 2.47E-39 | 19:45412955 | MeanAmyg |
| rs1516725 | Ventral_Striatum_(right) | 1.65E-38 | 3:185824004 | MeanHippo |
| rs1516725 | Supramarginal_Gyrus_(left) | 1.37E-31 | 3:185824004 | MeanHippo |
| rs2075650 | Supracalcarine_Cortex_(right) | 9.71E-29 | 19:45395619 | MeanAccur |
| rs2075650 | Supracalcarine_Cortex_(right) | 9.71E-29 | 19:45395619 | MeanHippo |
| rs1516725 | V_Cerebellum_(right) | 9.18E-27 | 3:185824004 | MeanHippo |
| rs4420638 | Supracalcarine_Cortex_(right) | 9.20E-08 | 19:45422946 | MeanAccur |
| rs4420638 | Supracalcarine_Cortex_(right) | 9.20E-08 | 19:45422946 | MeanHippo |
| rs4420638 | Supracalcarine_Cortex_(right) | 9.20E-08 | 19:45422946 | MeanThala |
| rs4420638 | Supracalcarine_Cortex_(right) | 9.20E-08 | 19:45422946 | MeanAmyg |
| rs429358 | Supracalcarine_Cortex_(right) | 1.78E-06 | 19:45411941 | MeanHippo |
| rs429358 | Supracalcarine_Cortex_(right) | 1.78E-06 | 19:45411941 | MeanAccur |
| rs429358 | Supracalcarine_Cortex_(right) | 1.78E-06 | 19:45411941 | MeanAmyg |
| rs429358 | Supracalcarine_Cortex_(right) | 1.78E-06 | 19:45411941 | MeanThala |
| rs1004787 | Amygdala_(left) | 7.24E-06 | 2:45159091 | MeanCaud |
| rs1004787 | Amygdala_(left) | 7.24E-06 | 2:45159091 | Mean_cun |
| rs1004787 | Amygdala_(left) | 7.24E-06 | 2:45159091 | Mean_late |
| rs557042 | Putamen_(right) | 2.94E-05 | 6:27845129 | Mean_Full_ |
| rs200965 | Putamen_(right) | 0.00012986 | 6:27866384 | Mean_cau |
| rs200965 | Putamen_(right) | 0.00012986 | 6:27866384 | Mean_Full_ |
| rs438613 | Juxtapositional_Lobule_Cortex_(right) | 0.0031296 | 3:28072086 | MeanAmyg |
| rs10801908 | Juxtapositional_Lobule_Cortex_(right) | 0.00838141 | 1:117090493 | Mean_insu |
| rs6859 | Supracalcarine_Cortex_(right) | 0.01411128 | 19:45382034 | MeanThala |
| rs6859 | Supracalcarine_Cortex_(right) | 0.01411128 | 19:45382034 | MeanHippo |
| rs6859 | Supracalcarine_Cortex_(right) | 0.01411128 | 19:45382034 | MeanAccur |
| rs157580 | Supracalcarine_Cortex_(right) | 0.01710901 | 19:45395266 | Mean_late |
| rs1516725 | Superior_Temporal_Gyrus,_(left) | 0.01944202 | 3:185824004 | MeanHippo |
| rs266058 | Amygdala_(left) | 0.02113597 | 2:104086428 | Mean_Full_ |
| rs266058 | Amygdala_(left) | 0.02113597 | 2:104086428 | Mean_isth |
| rs75627662 | Supracalcarine_Cortex_(right) | 0.02171833 | 19:45413576 | Mean_infe |
| rs28399637 | Supracalcarine_Cortex_(right) | 0.02409106 | 19:45324138 | MeanAmyg |
| rs1381287 | Amygdala_(left) | 0.0269796 | 14:98597552 | Mean_Full_ |

| SNPs | IDPs (Vol. of grey matter) | adj-p | POS | Enigma IDP |
|---|---|---|---|---|
| rs1081105 | Supracalcarine_Cortex_(right) | 4.26E-70 | 19:454129 | MeanAmygdala |
| rs1516725 | V_Cerebellum_(right) | 1.93E-64 | 3:1858240 | MeanHippocampus |
| rs1516725 | Supramarginal_Gyrus_(right) | 8.15E-64 | 3:1858240 | MeanHippocampus |
| rs993137 | Amygdala_(left) | 1.73E-54 | 3:8549903 | Mean_inferiortemporal_surfavg |
| rs993137 | Amygdala_(left) | 1.73E-54 | 3:8549903 | Mean_Full_Thickness |
| rs75627662 | Supracalcarine_Cortex_(right) | 2.75E-34 | 19:454135 | Mean_inferiortemporal_surfavg |
| rs1004787 | Amygdala_(left) | 6.21E-32 | 2:4515909 | MeanCaudate |
| rs1004787 | Amygdala_(left) | 6.21E-32 | 2:4515909 | Mean_lateraloccipital_surfavg |
| rs1004787 | Amygdala_(left) | 6.21E-32 | 2:4515909 | Mean_cuneus_surfavg |
| rs28399637 | Supracalcarine_Cortex_(right) | 2.77E-26 | 19:453241 | MeanAmygdala |
| rs429358 | Supracalcarine_Cortex_(right) | 4.95E-14 | 19:454119 | MeanHippocampus |
| rs429358 | Supracalcarine_Cortex_(right) | 4.95E-14 | 19:454119 | MeanAccumbens |
| rs429358 | Supracalcarine_Cortex_(right) | 4.95E-14 | 19:454119 | MeanAmygdala |
| rs429358 | Supracalcarine_Cortex_(right) | 4.95E-14 | 19:454119 | MeanThalamus |
| rs2075650 | Supracalcarine_Cortex_(right) | 6.18E-11 | 19:453956 | MeanHippocampus |
| rs2075650 | Supracalcarine_Cortex_(right) | 6.18E-11 | 19:453956 | MeanAccumbens |
| rs157580 | Supracalcarine_Cortex_(right) | 6.66E-07 | 19:453952 | Mean_lateraloccipital_surfavg |
| rs3888190 | Superior_Temporal_Gyrus_(left) | 1.14E-06 | 16:288894 | Mean_inferiortemporal_surfavg |
| rs3888190 | Superior_Temporal_Gyrus_(left) | 1.14E-06 | 16:288894 | MeanPutamen |
| rs3888190 | Superior_Temporal_Gyrus_(left) | 1.14E-06 | 16:288894 | MeanCaudate |
| rs3888190 | Superior_Temporal_Gyrus_(left) | 1.14E-06 | 16:288894 | MeanAccumbens |
| rs266058 | Amygdala_(left) | 0.000835 | 2:1040864 | Mean_isthmuscingulate_surfavg |
| rs266058 | Amygdala_(left) | 0.000835 | 2:1040864 | Mean_Full_Thickness |
| rs1568452 | Brain-Stem | 0.001911 | 2:5801283 | Mean_caudalanteriorcingulate_surfavg |
| rs1568452 | Brain-Stem | 0.001911 | 2:5801283 | Mean_caudalmiddlefrontal_surfavg |
| rs1568452 | Brain-Stem | 0.001911 | 2:5801283 | MeanAccumbens |
| rs1568452 | Brain-Stem | 0.001911 | 2:5801283 | Mean_isthmuscingulate_surfavg |
| rs1516725 | Ventral_Striatum_(right) | 0.004642 | 3:1858240 | MeanHippocampus |
| rs10119 | Supracalcarine_Cortex_(right) | 0.006895 | 19:454066 | MeanAccumbens |
| rs1516725 | Supramarginal_Gyrus_(left) | 0.01498 | 3:1858240 | MeanHippocampus |
| rs1516725 | Superior_Temporal_Gyrus_(left) | 0.016962 | 3:1858240 | MeanHippocampus |
| rs557042 | Putamen_(right) | 0.017487 | 6:2784512 | Mean_Full_Thickness |
